# PrimateAI-3D outperforms AlphaMissense in real-world cohorts

**DOI:** 10.1101/2024.01.12.24301193

**Authors:** David A. Parry, Tom Bosc, Tobias Hamp, Petko P. Fiziev, Abhishek Sharma, Irfahan Kassam, Jeremy McRae, Kyle Kai-How Farh

## Abstract

Accurately predicting the impact of genetic variants is essential for interpreting genomic data, yet no consensus exists on how to measure classifier performance. We prepared the most comprehensive set of benchmarks to date and applied them to the recently published models PrimateAI-3D and AlphaMissense. PrimateAI-3D outperforms AlphaMissense on rare-disease cohort and biobank benchmarks, indicating that performance on clinical databases or *in vitro* assays does not reliably generalize to real-world cohorts.

## Main

Genetic variants underlie many of the inter-individual differences in disease susceptibility in the human population, yet the clinical significance of most of the ∼70-million possible missense variants in the genome remain unknown. The scale of this problem has motivated the development of numerous computational algorithms for variant effect prediction, but there remains no agreed standard for benchmarking their accuracy, which hinders advancements in the field. Clinical labels from variant annotation databases, such as ClinVar and HGMD, are widely used as truth sets for model evaluation despite the acknowledgment of a range of biases and circularities leading to overoptimistic performance measures for models that may not generalize well to real-world data^1,2^. We previously developed a comprehensive set of six clinical benchmarks to evaluate the accuracy of PrimateAI-3D alongside other variant classifiers^3^, with a focus on performance in real-world cohorts. In addition to updating these previous benchmarks with the latest data, we introduce the largest benchmark to date, measuring the effects of genetic variants on 701 plasma proteins in participants from the UK Biobank Pharma Proteomics Project^4^, for a total of 29.3 million individual protein measurements. We use these benchmarks to compare the performance of PrimateAI-3D versus the recently published AlphaMissense tool, which was published after PrimateAI-3D, and hence has not been evaluated on comprehensive real-world cohorts.

First, we compared PrimateAI-3D with AlphaMissense on updated versions of the six benchmarks which had previously been used to benchmark PrimateAI-3D and 16 other computational algorithms in Gao *et al*.^3^ (Methods). PrimateAI-3D had been shown to outperform each of the other algorithms in all six benchmarks, but had not been benchmarked against AlphaMissense, which was published after PrimateAI-3D and only included a limited set of benchmarks. We found that PrimateAI-3D outperformed AlphaMissense in all four large clinical cohorts consisting of half a million individuals (DDD, ASD, CHD and UKBB, Figure 1 and Table S1). Although the AlphaMissense paper stated that they used clinically annotated variants from the ClinVar database for model selection and hyperparameter optimization, and evaluation data to select the optimal training iteration (including saturation mutagenesis data from ProteinGym), we did not attempt to remove variants that had previously been utilized by AlphaMissense. For the 31 genes tested with *in vitro* saturation mutagenesis assays, AlphaMissense performed better in 16 genes, while PrimateAI-3D performed better in 15 of the genes. AlphaMissense also performed slightly better on the ClinVar benchmark (mean AUC = 0.951 vs 0.940).

**Figure 1.**
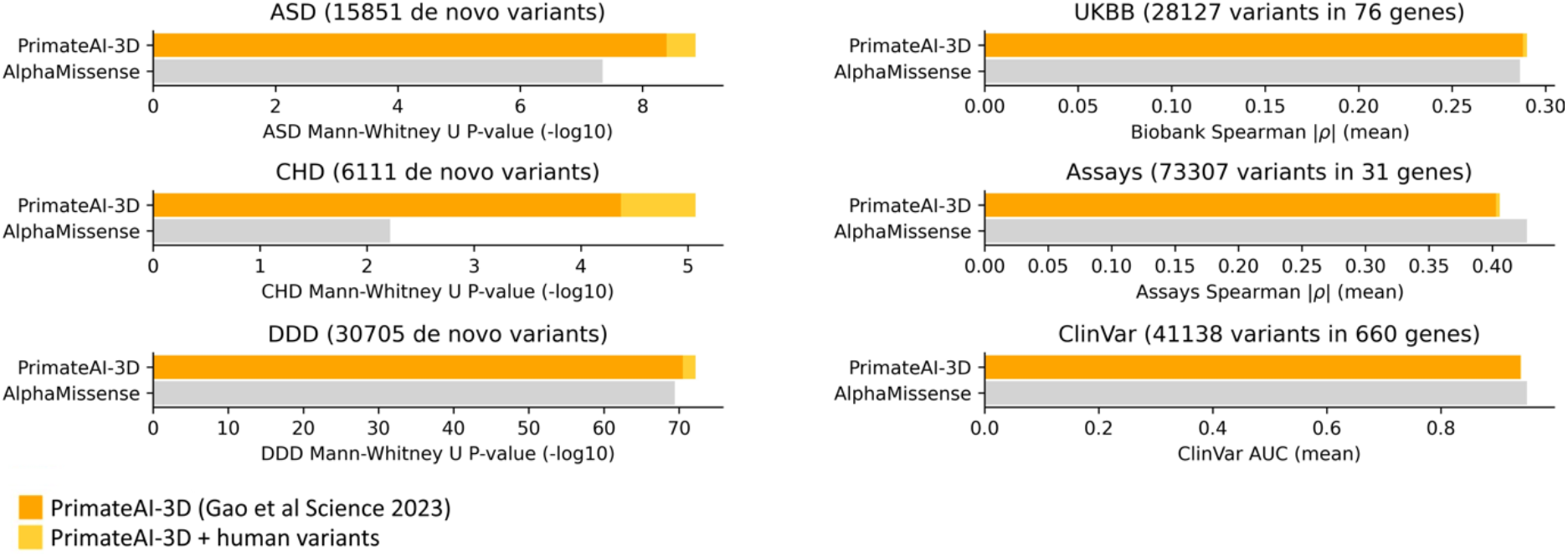
PrimateAI-3D versus AlphaMissense performance on six previously published benchmarks. Performance for AlphaMissense (gray), published PrimateAI-3D scores (dark orange), and updated PrimateAI-3D scores to include training on human variant data (light orange) are shown.

We devised a new proteomics benchmark from the UK Biobank where we correlated classifier scores with decreases in protein levels detected in blood plasma (Methods), as pathogenic protein-coding variants frequently decrease protein levels^5^. This comprises the largest ever real-world benchmarking dataset with combined protein measurements and sequence data from 41,836 individuals across 701 genes, with 29.3 million unique protein observations. Because this benchmark had not been previously tested, we assessed the performance of 16 other classifiers alongside PrimateAI-3D and AlphaMissense (Figure 2A, Table S2). PrimateAI-3D showed the highest mean inverse correlation with protein levels out of all methods tested. Notably, the top 4 computational predictors were all deep learning models.

**Figure 2.**
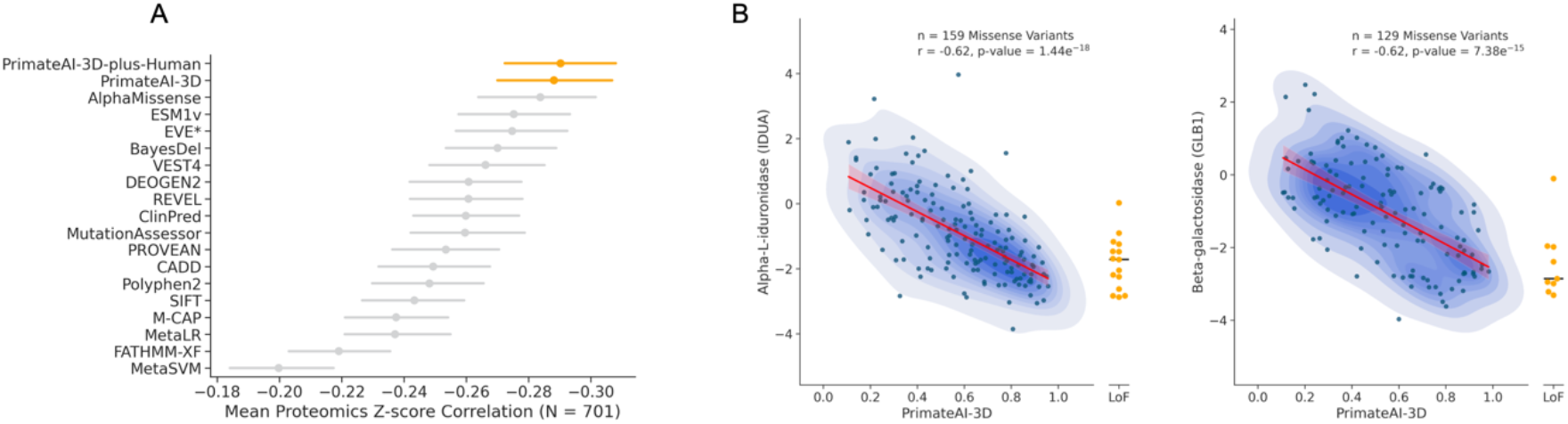
Classifier performance on a new UK Biobank proteomics benchmark. (A) Correlation between protein levels in blood plasma and classifier score. (B) Example PrimateAI-3D score correlations with disease-associated proteins IDUA and GLB1.

AlphaMissense shares many similarities with PrimateAI-3D, such as adopting PrimateAI’s method of training the model using common primate variants as benign labels versus mutation rate-matched unobserved variants as pathogenic labels, and utilizing 3-D structural information and protein language models. Hence, the two scores are highly correlated (Pearson correlation = 0.83, Figure S1), partially explaining their close performance in the proteomics benchmark. A notable difference between the two methods is that AlphaMissense uses predominantly human missense variants for training while most of PrimateAI-3D’s training data comes from non-human primate sequencing. We therefore decided to assess the performance of PrimateAI-3D when trained exclusively with human variant data and ensembled with the model trained with predominantly primate data (Methods). This new version, “PrimateAI-3D-plus-human” demonstrated improved performance on all seven benchmarks (Figure 1, 2A) indicating that the model can learn from both primate and human variant datasets. Further work is needed to investigate the complementary nature of these variant training sets and may provide novel insights into features discriminating benign from pathogenic variation.

Herein we describe the most comprehensive set of real-world cohorts for benchmarking missense variant classification to date, tabulating the effects of variants in nearly half a million individuals, which we provide to the community as a resource to further accelerate the development of computational tools. Importantly, these benchmarking cohorts are independent of human labels and their inherent biases. We find that PrimateAI-3D outperforms AlphaMissense in all real-world cohorts, including on both biobank benchmarks, which directly assess variant impacts on clinical phenotypes and blood protein levels, and on three rare disease cohorts, which evaluate the classifier’s ability to distinguish *de novo* mutations observed in disease patients compared to a healthy control population. Although AlphaMissense did not do as well in real-world cohorts, it performed best on clinically annotated variants from ClinVar and *in vitro* saturation mutagenesis assays, which may be explained by its use of ClinVar for model selection and hyperparameter tuning during training and use of evaluation data for early stopping. These two datasets also display higher evolutionary conservation than the genome wide average (Figure S2) suggesting that optimal performance on these benchmarks may not generalize to the rest of the proteome where evolutionary signals are not so strong. Our results show the importance of testing variant classifiers on real-world cohorts when assessing their clinical utility. It also emphasizes the need to benchmark scores on a wide range of datasets to ensure scores do not overfit to specific use-cases.

Notably, for UK Biobank clinical phenotype^3^ and proteomics benchmarks, both based on direct measurements and not influenced by manually curated clinical labels, deep-learning models all outperform more basic classifiers. The superior performance of these models on these benchmarks suggests that deep-learning strategies are better able to identify biologically relevant effects of missense variation and are a leading avenue for future developments.

## Data Availability

Benchmark data used in this study will be made available upon publication.

## Methods

### De novo Mutation Benchmarks

We gathered de novo mutations (DNMs) from published studies of developmental disorders (DDD)^6–9^, autism spectrum disorders (ASD)^10–18^ and congenital heart disorders (CHD)^19^. DDD had 27,030 DNMs from 18,066 patients, ASD 11,824 DNMs from 8,089 patients and CHD 1,855 DNMs from 1,350 patients. Each cohort was compared against 4,389 DNMs from 2,997 healthy controls gathered from multiple studies^11–13,15,17,18,20^ using the Mann-Whitney U test to evaluate how well each classifier can distinguish missense DNMs observed in disease cohorts from those in healthy controls.

### ClinVar Benchmark

Missense variants labelled as either benign, likely benign, likely pathogenic or pathogenic and with at least a 1-star review status were selected from ClinVar (downloaded September 19^th^ 2021). The ability of each classifier to discriminate between benign/likely benign and pathogenic/likely pathogenic variants was determined per-gene using the area under the receiver operating characteristic curve (AUC). Genes were limited to those with at least 5 (likely) pathogenic and 5 (likely) benign variants scored by both classifiers.

### Deep Mutational Scan Benchmark

We downloaded deep mutagenesis assays for human proteins from the September 2022 versions of ProteinGym^21^ and MaveDB^22^ (89 assays). We filtered assays so that any two assays from the same gene had <80% overlap in variants or <80% correlation in assay score (52 assays). For each protein, we then only kept the assay with highest average correlation to PrimateAI-3D and AlphaMissense (31 assays and genes). Model performance was evaluated by computing the absolute Spearman rank correlation between model prediction scores and assay scores for each assay and then taking the mean across all assays.

### UK Biobank Clinical Phenotyping Benchmark

The use of UK Biobank data was approved by the UK Biobank under application no. 33751. The UK Biobank clinical phenotyping benchmark used data from 454,712 individuals with both exome sequencing data and broad clinical phenotyping from the UK Biobank. This dataset consists of gene-phenotype pairs where presence of rare protein-coding variants (protein-truncating or missense) is significantly associated with a quantitative phenotype on burden testing (P < 10^-10^), without stratifying missense variants by pathogenicity. When a gene was associated with multiple phenotypes the phenotype with the most significant association was chosen. In total 76 gene-phenotype pairs were used consisting of phenotype data for 33 unique traits. Quantitative traits were standardized by inverse rank normal-transformation and model performance evaluated by computing the absolute Spearman rank correlation between model prediction scores and mean standardized phenotype scores for carriers of variants in each gene. The mean per-gene correlations were compared between tools.

### UK Biobank Plasma Proteome Benchmark

We utilized data for plasma protein levels of 1,453 proteins in conjunction with whole exome sequencing in 41,836 unrelated individuals of European descent from the UK Biobank. We selected a subset of 829 genes where protein truncating variants had a significant negative effect on plasma levels in the rare variant burden test at Bonferroni corrected p-value threshold of 5% after testing 1,453 gene-protein pairs. In Figure 2A, we additionally restricted the evaluation to missense variants that are scored by all classifiers and genes with at least 10 such variants, which reduced the number of genes to 701 with 42,409 variants assessed in total. Plasma protein levels were corrected for a range of covariates including age, sex, ancestry and medication use as described previously^23^. Subsequently, plasma levels were adjusted for the top 20 proteomics principal components and inverse rank normal transformed. Spearman correlation was computed on variant level between pathogenicity scores and the average plasma levels across all carriers.

### Training PrimateAI-3D with Additional Human Variant Data

We trained PrimateAI-3D as described previously^3^ but modified one of the three training objectives so that instead of differentiating common primate and human variants (>0.1% AF) from unobserved variants it was tasked with separating only human variants with a minimum allele frequency of 0.001% from unobserved variants. Ensembled scores were generated by averaging the ranks of published PrimateAI-3D scores and the new human-only trained model scores.

## Competing interests

All authors are employees of Illumina, Inc.

## Supplementary Information

**Figure S1.**
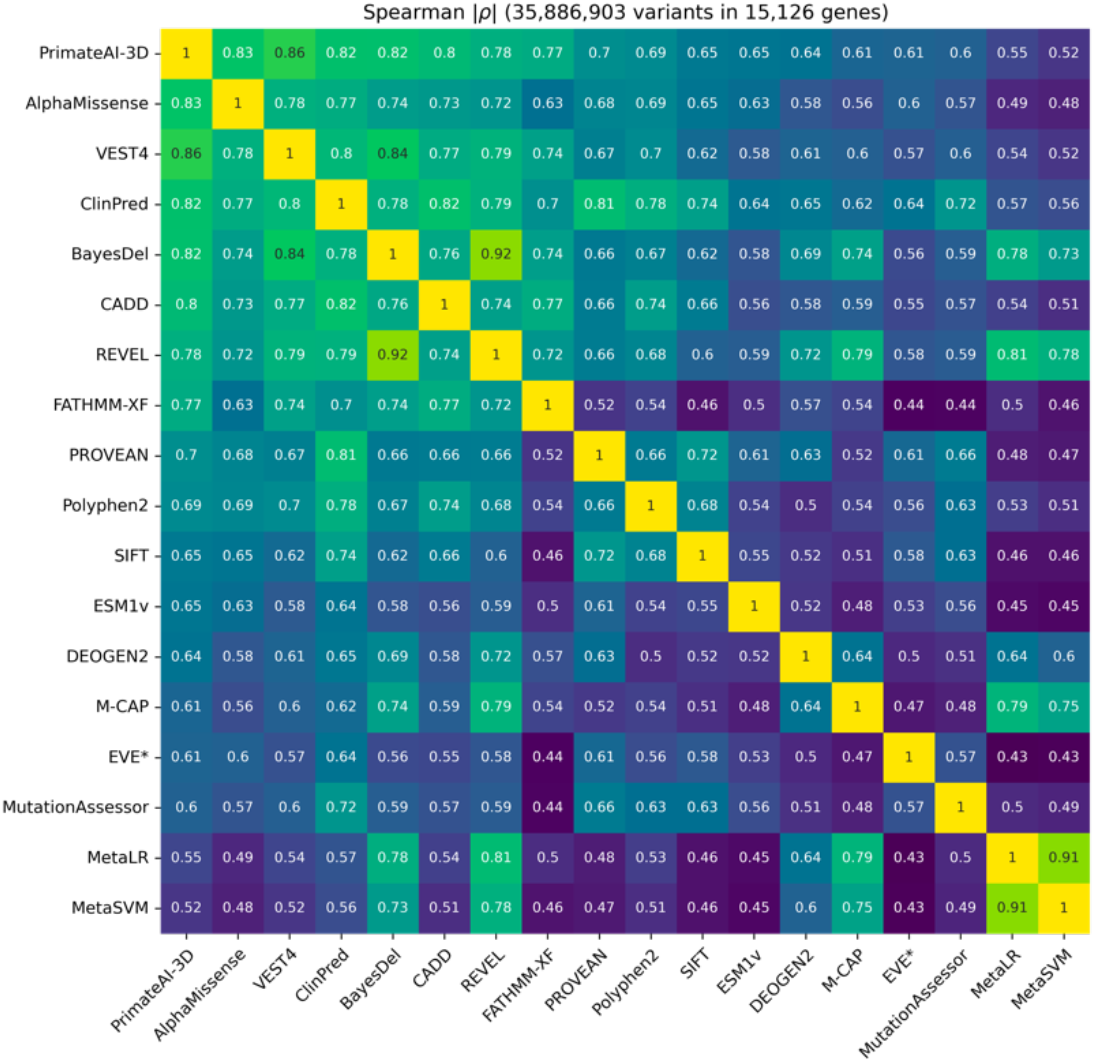
Pairwise correlations of classifier scores.

**Figure S2:**
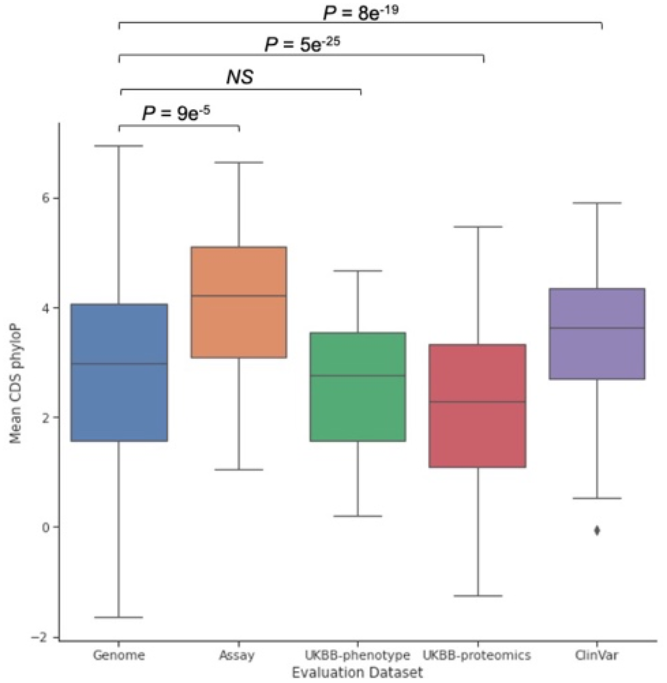
Evolutionary conservation of benchmark genes. The mean phyloP score from 100 vertebrates was calculated for the coding region of each gene either genome-wide or for each gene in the corresponding benchmark dataset. P-values are from Mann-Whitney U tests comparing mean coding region phyloP scores for genes in each evaluation dataset versus all genes.

**Table S1.**
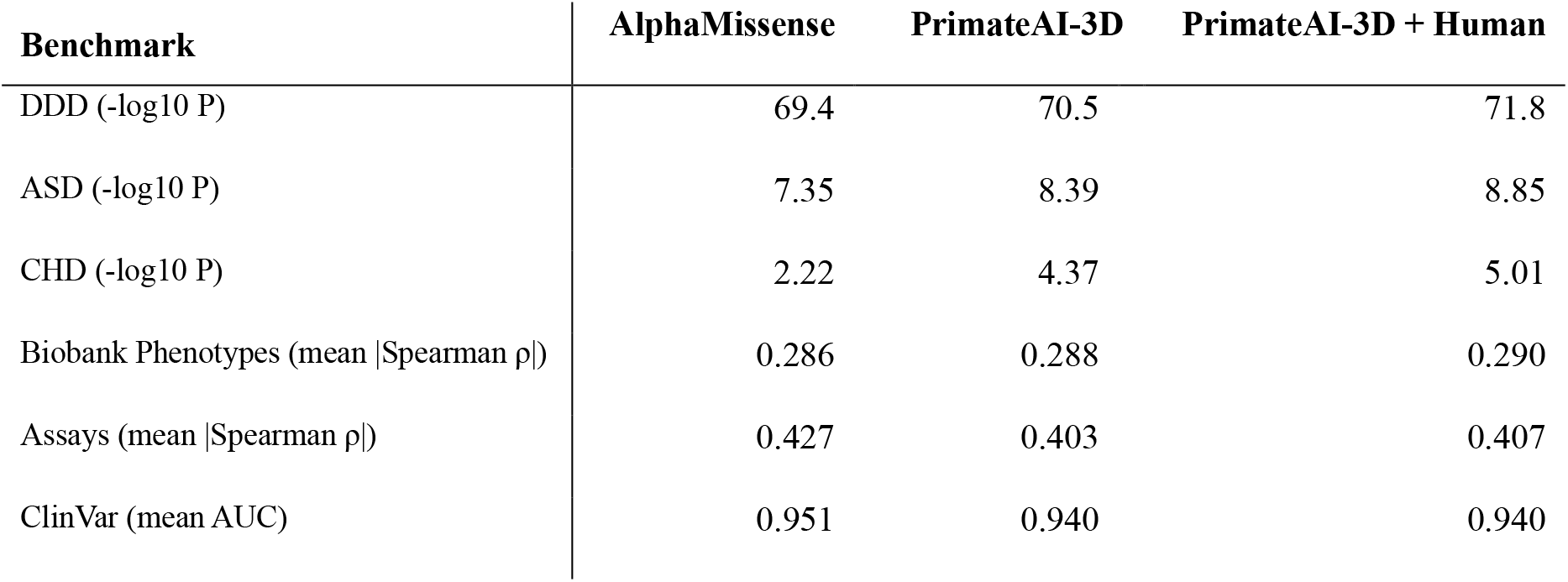
PrimateAI-3D and AlphaMissense comparisons on six benchmarks.

**Table S2.** PrimateAI-3D comparison against 17 pathogenicity classifiers on proteomics data from the UK Biobank.

| Score | Mean Proteomics Z-score<br>Spearman Correlation | Standard Error |
| --- | --- | --- |
| **PrimateAI-3D-plus-Human** | -0.290278 | 0.009071 |
| **PrimateAI-3D** | -0.288154 | 0.009054 |
| **AlphaMissense** | -0.283781 | 0.00939 |
| **ESM1v** | -0.275245 | 0.009096 |
| **EVE\*** | -0.274739 | 0.009346 |
| **BayesDel** | -0.270052 | 0.008828 |
| **VEST4** | -0.266177 | 0.009064 |
| **DEOGEN2** | -0.260764 | 0.008891 |
| **REVEL** | -0.260702 | 0.008995 |
| **ClinPred** | -0.259851 | 0.008688 |
| **MutationAssessor** | -0.259603 | 0.0089 |
| **PROVEAN** | -0.253448 | 0.008589 |
| **CADD** | -0.249388 | 0.008949 |
| **Polyphen2** | -0.248185 | 0.008844 |
| **SIFT** | -0.243338 | 0.00852 |
| **M-CAP** | -0.237486 | 0.008658 |
| **MetaLR** | -0.237139 | 0.008686 |
| **FATHMM-XF** | -0.21911 | 0.008674 |
| **MetaSVM** | -0.199805 | 0.008708 |

## Notes

### Funding Statement

This study did not receive any funding

### Author Declarations

Use of human data from UK Biobank was approved by the UK Biobank under application no. 33751. All other human data were obtained from datasets available to the public.

